# The Relationship Between Immune Cells and Heart Failure: A Study Integrating Immune Infiltration Analysis and Bidirectional Mendelian Randomization

**DOI:** 10.1101/2024.04.26.24306471

**Authors:** Yiding Yu, Huajing Yuan, Wenwen Liu, Yitao Xue, Xiujuan Liu, Wangjun Hou, Yan Li

## Abstract

In recent years, attempts have been made to employ various immunomodulatory therapies as new treatment strategies for heart failure, yet the outcomes have been disappointing. Consequently, unveiling the complexity and causal relationships of the immune system’s role in heart failure has become imperative.

**Methods:** We first determined the levels of immune cells in failing myocardium using five immune infiltration algorithms: Cibersort, xCell, ssGSEA, MCPcounter, and QuanTIseq. Subsequently, two-sample bidirectional Mendelian Randomization (MR) analysis was conducted between heart failure and 731 immune cell phenotypes using Inverse Variance Weighted (IVW) as the main method, reinforced by MR-Egger, Weighted median, and Weighted mode as secondary analysis methods. Sensitivity analyses were performed to ensure data stability and feasibility.

**Results:** Immune infiltration analysis revealed changes in the levels of immune cell infiltration in failing myocardium, including T cells, Mast cells, B cells, NK cells, and macrophages. MR analysis indicated that 19 immune cell phenotypes from 5 cell types were associated with an increased risk of heart failure, while 12 immune cell phenotypes from 6 cell types were associated with a decreased risk, without heterogeneity and pleiotropy. Reverse MR analysis did not support a causal relationship between HF and any of the 731 immune cell phenotypes.

**Conclusion:** Our study highlights the complex pattern and causal relationship between the immune system and heart failure through immune infiltration and MR analyses. This provides new insights for the exploration of immunocyte therapies for heart failure.

## INTRODUCTION

Cardiovascular diseases are among the leading causes of mortality worldwide, encompassing conditions such as hypertension, arrhythmias, coronary artery disease, heart failure, and other heart-related diseases. Heart failure, in particular, often represents the final stage of most cardiovascular diseases. It generally results from structural modifications or functional disorders of the heart, leading to impairments in ventricular filling or ejection capabilities^1^. As the population ages, the survival rates of ischemic heart disease improve, and more effective evidence-based therapies emerge, the prevalence of heart failure continues to rise, affecting over 64 million individuals globally^2^. Despite ongoing efforts to treat and manage heart failure, there has been little reduction in the burden of mortality and hospitalization^3^. Therefore, researching and identifying modifiable risk factors is crucial for the management and prevention of heart failure.

Heart failure is a complex clinical syndrome influenced by numerous factors, including cardiomyopathy, activation of the sympathetic nervous system, abnormalities in energy supply, cardiotoxic drugs, loss of cardiomyocytes, dysregulation of calcium handling, and epigenetic mechanisms^4–10^. In addition to these factors, in recent years, there has been an increasing recognition of the role of inflammation and the immune system in the pathophysiology of heart failure. Following myocardial injury or infection, a substantial influx of immune cells is recruited to the heart to clear dead tissues, eliminate pathogens, and facilitate healing^11^. Different subtypes of immune cells participate in the inflammatory response, repair processes, and cardiac remodeling in their unique ways, influencing the progression of heart failure. M2 macrophages, which exhibit anti-inflammatory functions, are primarily activated by the inflammatory cytokine IL-4. They mainly exert their effects by secreting anti-inflammatory cytokines such as IL-10 to inhibit M1 macrophages, thereby improving the function of the damaged heart and reducing cardiac fibrosis^12^.Depletion of CD8+ T lymphocytes can reduce apoptosis within ischemic myocardial tissue, hinder the inflammatory response, limit myocardial damage, and improve cardiac function^13^. CD4+ T cells, on the other hand, can modulate immune responses by secreting cytokines such as interferon-gamma (IFN-γ) and tumor necrosis factor-alpha (TNF-α), which promote inflammation and myocardial damage^14^. B cells are capable of regulating the heart’s adaptation to injury and ultimately influence the extent of cardiac functional impairment following damage^15^.Regulatory T cells (Tregs) are beneficial to the heart as they suppress excessive inflammatory responses in the early stages of heart damage and promote stable scar formation. However, in chronic heart failure, the phenotype and function of Tregs can shift towards a pro-fibrotic and anti-angiogenic cell type. This functional transformation has rekindled interest^16^. Additionally, various subtypes of immune cells, including neutrophils, dendritic cells, and NK cells, are involved to varying degrees in the development of heart failure^17–19^. Despite mounting evidence indicating a close connection between various subtypes of immune cells and heart failure, the outcomes of attempts at new treatment strategies using immune modulation therapies have been disappointing^20^. Therefore, unraveling the complexity of the role played by the immune system in heart failure, as well as its causal relationships, becomes critically important. In this study, we will utilize five immune infiltration algorithms to uncover the quantity of immune and stromal cells within the failing myocardium, illustrating the myocardium’s complex landscape of cellular heterogeneity. Moreover, through Mendelian Randomization (MR) analysis, we will examine the intricate causal and reverse causal relationships between 731 immune cell phenotypes of seven types of immune cells and heart failure. Our research aims to shed new light on the role of immune cells in the pathophysiology of heart failure and contribute to the development of immunomodulatory therapies. The outline of the design of our study was shown in Figure 1.

**Figure 1:**
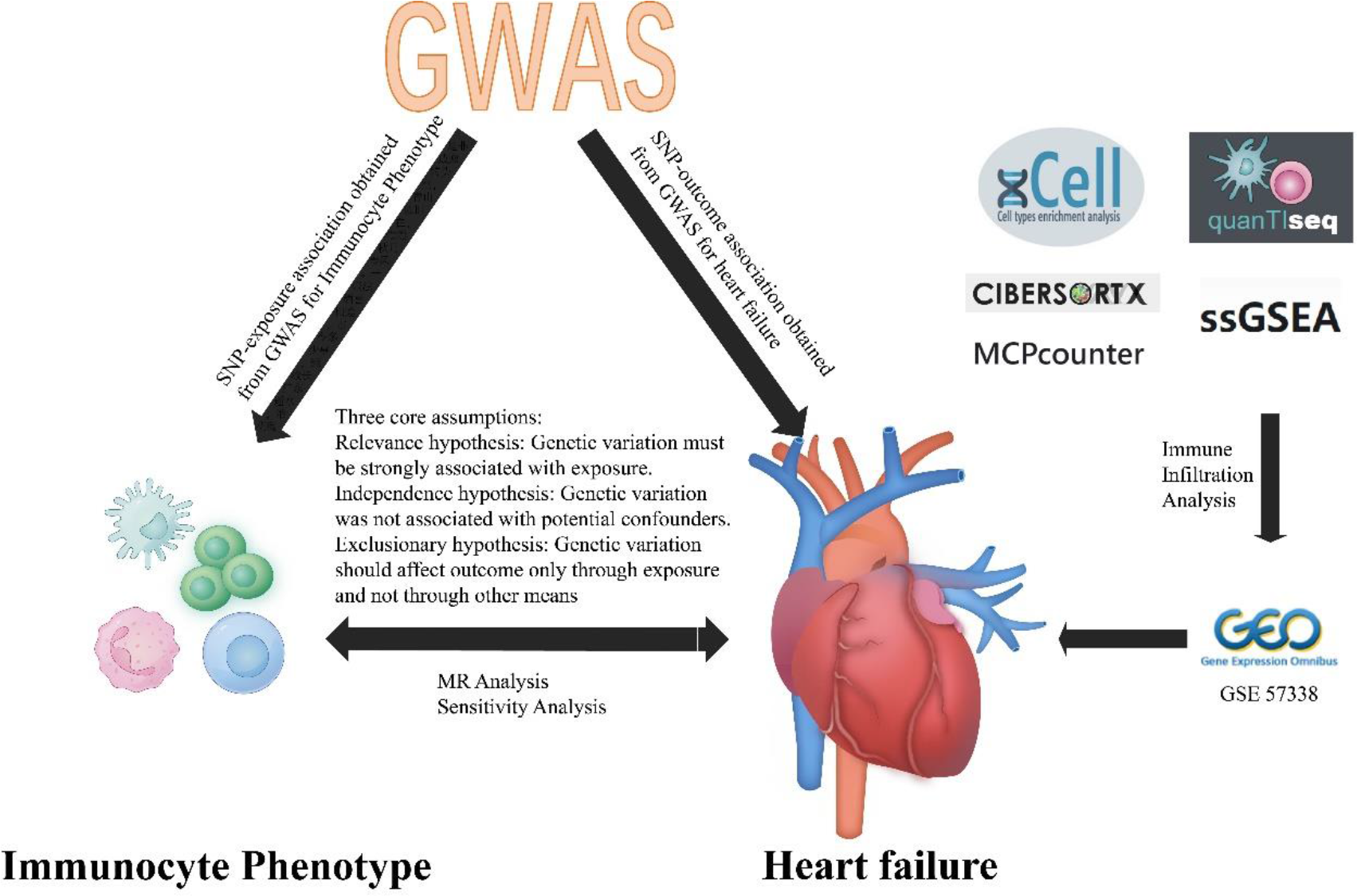
The study design.

## MATERIALS AND METHODS

### Immune Infiltration Analysis

To portray the intricate cell heterogeneity landscape of failing myocardium, we employ five immune infiltration algorithms: Cibersort, xCell, ssGSEA, MCPcounter, and QuanTIseq. The dataset we use is GSE57338, which consists of left ventricular myocardial samples from 95 patients with ischemic heart failure and 136 normal individuals. It represents the largest dataset of myocardial samples from heart failure patients currently available^21^. The Cibersort algorithm performs deconvolution analysis based on the principle of linear support vector regression, cataloging gene expression profiles of 22 types of immune cells. This includes seven types of T cells, naive and memory B cells, plasma cells, NK cells, and myeloid subsets^22^. The xCell algorithm employs ssGSEA to calculate enrichment scores for each cell type signature, enabling the assessment of infiltration levels for up to 64 cell types. This encompasses various adaptive and innate immune cells, hematopoietic stem cells, epithelial cells, and extracellular matrix cells^23^. The ssGSEA algorithm calculates immune cell infiltration based on markers for 24 types of immune cells provided by Bindea^24^. The MCPcounter algorithm uses a linear model to translate the expression levels of immune-related genes into the abundance of eight immune cells and two stromal cells, including T cells, CD8+ T cells, cytotoxic lymphocytes, NK cells, B lymphocytes, monocytes, myeloid dendritic cells, neutrophils, as well as endothelial cells and fibroblasts^25^. The quanTIseq method utilizes a deconvolution algorithm to predict the composition of various types of immune cells within a sample, including 10 immune cell types such as B cells, Monocytes, Neutrophils, Natural killer (NK) cells, and Non-regulatory CD4+ T cells^26^. Cell distribution differences between the heart failure group and the normal group are compared using the t-test, with the threshold set at an adjusted p-value of less than 0.05.

### MR Study design

In our study, we evaluated the causal relationships between 731 immune cell phenotypes (grouped into 7 categories) and heart failure using bidirectional two-sample Mendelian Randomization (MR) analysis. Our research adheres to the three core assumptions of MR analysis: 1. Genetic variants must be strongly associated with the exposure. 2. Genetic variants are uncorrelated with potential confounding factors. 3. Genetic variants should only influence the outcome through the exposure and not through other pathways^27^. All data utilized in this study are publicly available and have been obtained with the consent of the relevant participants and ethical approval, thus institutional review board ethical approval was not required for this research.

### Exposure and Outcome Data Acquisition

The GWAS catalog (GCST90001391 to GCST90002121) provides a summary of GWAS statistical data for each immunological trait^28^. It encompasses integrated data collected from 3,757 individuals of European ancestry, covering 731 immune phenotypes. As for the outcomes, we selected heart failure data from the HERMES consortium, which includes 47,309 heart failure patients and 930,014 controls^29^.

### IVs selection

During the selection process for SNPs, we encountered challenges in identifying effective SNPs due to the stringent genome-wide significance threshold (P < 5E-8) applied in our GWAS screening. Consequently, we ultimately opted for a threshold where SNP selection satisfied P < 1E-5^30^. To prevent linkage disequilibrium (LD) from affecting the analysis results, we set the LD threshold parameter r to 0.001 and used a distance of 10,000 Kb for SNP analysis. Subsequently, we used the PhenoScanner V2 database, a human genotype-phenotype association repository, to further verify the inclusion of SNP loci and to check for any other potential confounding variables associated with the included SNP loci. Lastly, to assess whether the included SNPs were affected by weak instrumental variables, we excluded values with an F-statistic greater than 10 (calculated using the formula F = β^2 / SE^2, where β represents the allele effect size, and SE is the standard error). If the F-statistic for SNPs is less than 10, it indicates the possibility of a weak instrument variable bias with those SNPs, and therefore, they are excluded to avoid affecting the results. Subsequently, we extract outcome information through the IEU OpenGWAS database and ascertain the relationships among SNPs that meet the assumed criteria from the results. By merging the exposure data set with the generated data set and removing duplicate sequences, we obtain eligible instrumental variables (IVs) that meet the standards for further analysis to establish genetic associations.

### Statistical analysis

In our study, Mendelian Randomization (MR) analysis was conducted using the TwoSampleMR and MR-PRESSO packages within the R software, version 4.2.0. In our primary analysis, the Inverse Variance Weighted (IVW) model was applied to assess the causal relationship between each immune cell phenotype and the risk of heart failure ^31^. The characteristic of the IVW method is that it does not consider the presence of an intercept and uses the inverse of the final variance (the square of standard error) as the weight for fitting. When using IVW to explore causal relationships, there may be other factors affecting the bias estimation of genetic diversity and causal effects. MR-Egger regression can detect horizontal pleiotropy through the p-value of the intercept. The principle is to check whether the outcome effect is zero when the instrumental variable effect is zero (i.e., whether the intercept is zero). If the intercept is not zero, it indicates the existence of horizontal pleiotropy, which implies that the outcome exists even without exposure^32^. The MR-PRESSO method is capable of detecting outlier SNPs and, assuming that the SNPs utilized are valid, provides a causal estimate after the potential outliers are removed^33^. Due to the diversity of study subjects and experimental conditions, two-sample Mendelian Randomization (MR) analysis may exhibit heterogeneity, leading to a potential bias in the estimation of causal effects. We primarily assess heterogeneity using Cochran’s Q statistic, with a p-value less than 0.05 indicating significant heterogeneity. When using the random-effects IVW as the primary outcome measure, heterogeneity is considered acceptable. If the p-value is greater than 0.05 in the test, there is no evidence of heterogeneity among the included instrumental variables (IVs), meaning that the impact of heterogeneity on the causal effect estimation can be ignored. Finally, to evaluate the influence of individual SNPs on the overall estimate, a “leave-one-out” analysis is conducted by sequentially excluding each SNP from the analysis. The leave-one-out sensitivity test calculates the MR results of the remaining IVs after each IV is removed one by one. If the MR results estimated by the other IVs differ significantly from the overall result after the exclusion of a particular IV, this indicates that the MR results are sensitive to that IV. If there is no significant change in the overall results, the results are considered reliable. All results are presented as odds ratios (OR) with 95% confidence intervals (CI), and when p<0.05, the results are considered to be statistically significant.

## RESULTS

### Immune Infiltration Analysis of Myocardium

We conducted an immune infiltration analysis on the dataset GSE57338 utilizing five immunoinfiltration algorithms: Cibersort, xCell, ssGSEA, MCPcounter, and QuanTIseq. According to the Cibersort algorithm, differences were observed in 6 immune cell types. The xCell algorithm indicated differences in 24 cell types. The ssGSEA algorithm revealed discrepancies in 15 immune cell types. The MCPcounter algorithm showed variances in 2 immune cells and 1 stromal cell type. The QuanTIseq algorithm demonstrated differences in 6 immune cell types. The results of the analysis and cell expression are displayed in Figure 2.

**Figure 2:**
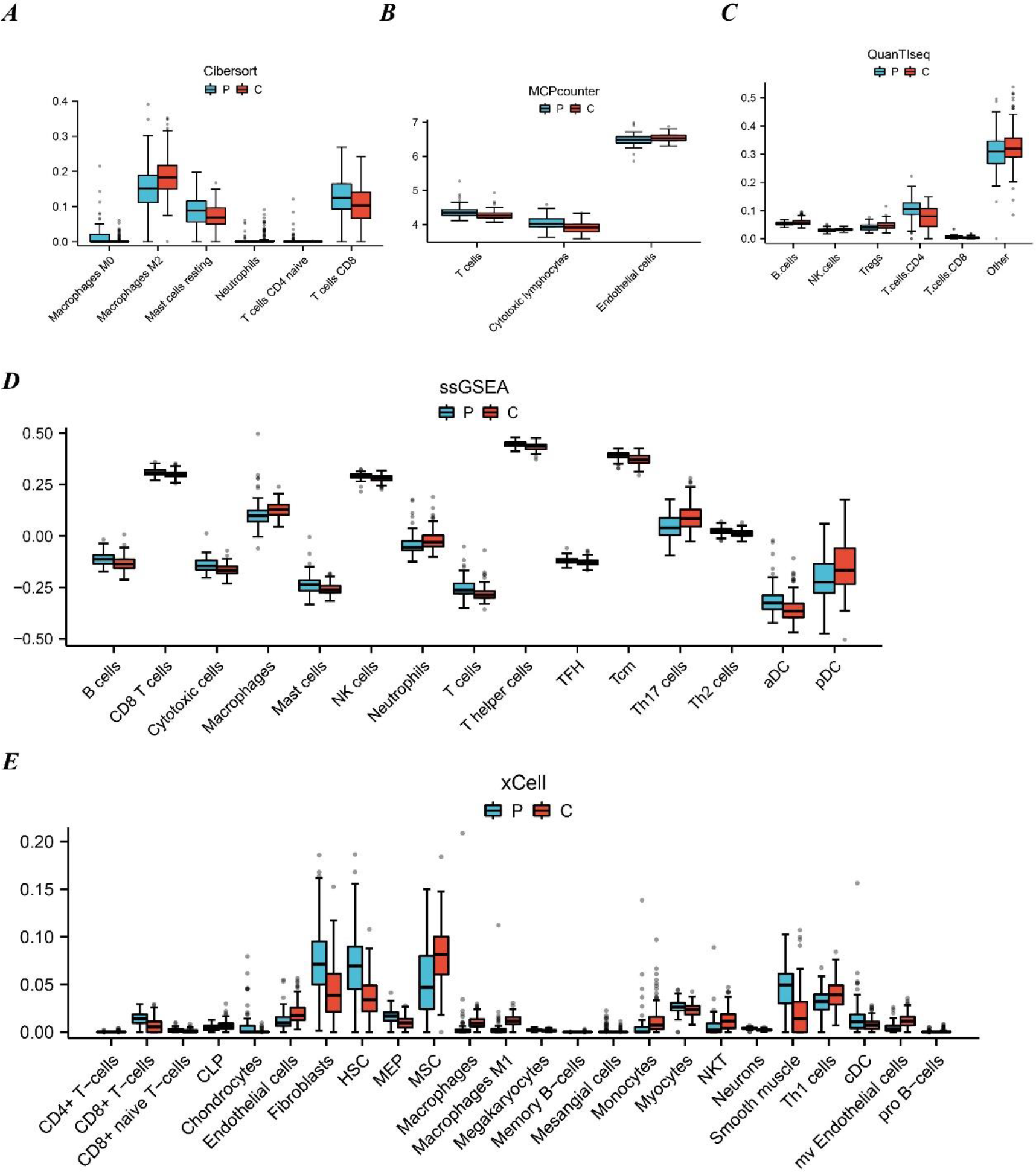
Immune cell infiltration analysis between HF and control. (A) Result of Cibersort algorithm. (B) Result of MCPcounter algorithm. (C) Result of QuanTIseq algorithm. (D) Result of ssGSEA algorithm. (E) Result of xCell algorithm.

### Effect of Immunocyte on HF

We conducted instrumental variable (IV) selection on GWAS data for 731 immune cell phenotypes, with all IVs having F values greater than 10, indicating no presence of weak instrument bias. The results from the Inverse Variance Weighted (IVW) method predicting genetic instruments for immune cell phenotypes with causal relationships to heart failure are shown in Figure 3. Our analysis indicates that 19 immune cell phenotypes across 5 immune cell groups are associated with an increased risk of heart failure (OR>1, P<0.05). Specifically, B cell phenotypes include: CD19 on IgD+, IgD+ AC, IgD on IgD+ CD38dim, CD27 on IgD- CD38dim, IgD on IgD+ CD24+, CD27 on IgD- CD38-, IgD+ CD38br AC, CD38 on IgD+ CD38br, CD19 on IgD+ CD38br, Sw mem %lymphocyte, and BAFF-R on IgD- CD38dim. cDC phenotype: CCR2 on CD62L+ myeloid DC. Myeloid cell phenotype: CD45 on lymphocyte. TBNK phenotype: TCRgd %lymphocyte. Treg phenotypes include: CD28+ CD45RA+ CD8br %T cell, CD28+ CD45RA+ CD8br AC, CD39+ CD8br %CD8br, CD28- CD127- CD25++ CD8br %CD8br, and CD28- CD127- CD25++ CD8br %T cell. There are 12 immune cell phenotypes across 6 immune cell groups that are associated with a decreased risk of heart failure (OR<1, P<0.05). Specifically, B cell phenotypes include: CD20 on IgD+ CD38br, Unsw Mem %lymphocyte, BAFF-R on IgD+ CD38br, and CD20 on IgD- CD38dim. cDC phenotype: CD62L- DC %DC. Maturation stages of T cell phenotype: HVEM on TD CD4+. Myeloid cell phenotype: CD11b on CD14+ monocyte. TBNK phenotypes include: HLA DR++ monocyte %leukocyte, SSC-A on HLA DR+ NK, SSC-A on CD14+ monocyte, and CD45 on CD8br. Treg phenotype: Activated Treg AC. Results for all analytical methods are provided in Supplementary File 1.

**Figure 3:**
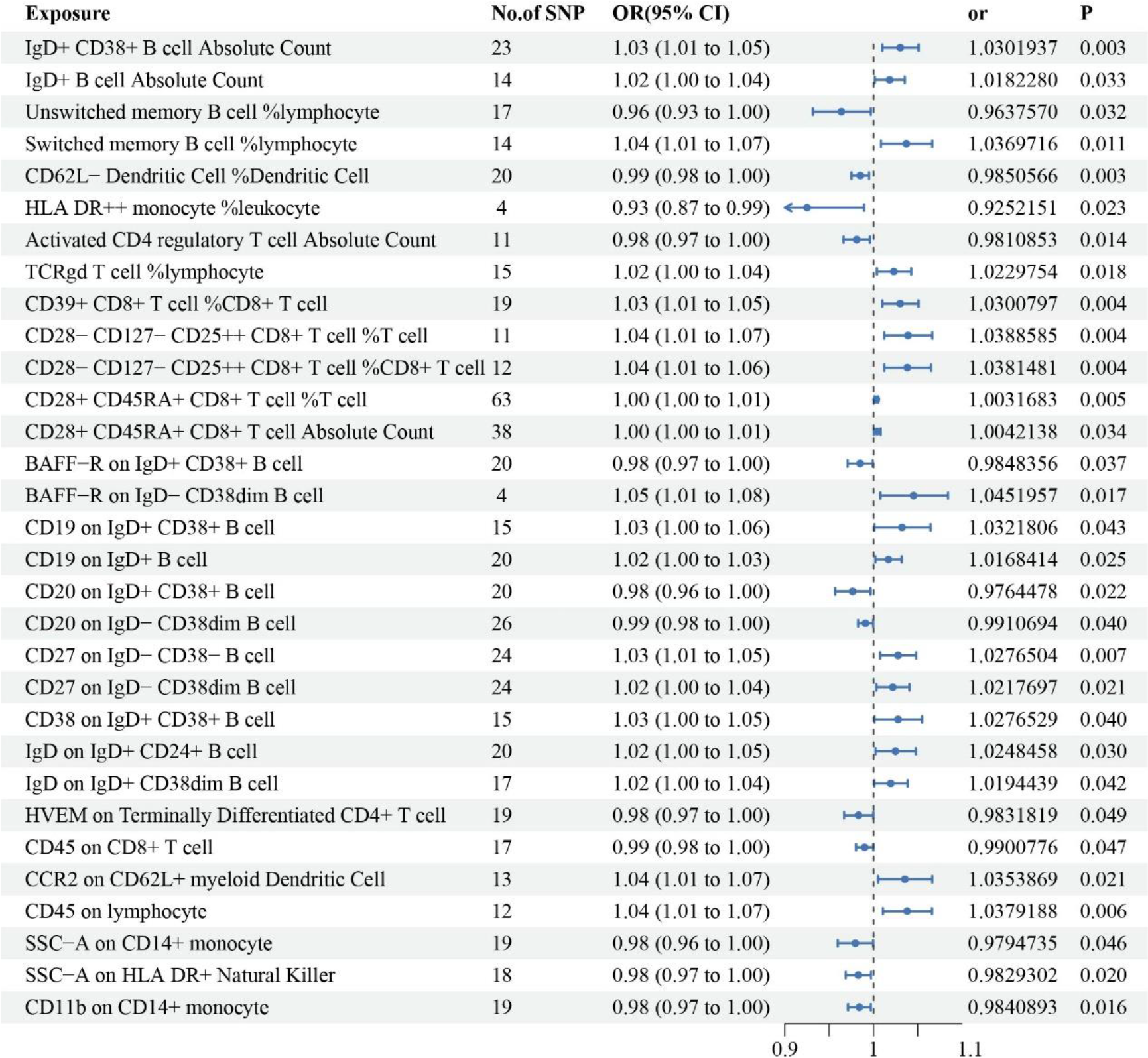
Associations between immune cell phenotypes and risk of heart failure.

### Sensitivity Analysis of the Effect of Immunocyte on HF

The sensitivity analysis results indicate that the immune cell phenotypes used for MR analysis in heart failure do not present evidence of heterogeneity (Q test P>0.05), horizontal pleiotropy (MR-Egger intercept method P>0.05), or significant outliers, suggesting that our findings are robust and credible. The “leave-one-out” analysis reveals that sequentially excluding each single nucleotide polymorphism (SNP) yields results from the remaining SNPs’ IVW analysis that are consistent with the outcomes obtained when all SNPs are included, therefore indicating that no single SNP has a substantial effect on the estimated causal association. The results of heterogeneity and pleiotropy analysis are shown in Supplementary file 2.

### Effect of HF on Immunocyte

We performed IV selection on the GWAS data for HF, ensuring that all IVs had an F statistic greater than 10 to avoid weak instrument bias. Unfortunately, our MR analysis did not uncover any causal relationships between HF and the 731 immune cell phenotypes examined. As such, we found no bidirectional causality between immune cell phenotypes and heart failure.

## DISCUSSION

In our study, we initially employed five different immune infiltration analysis algorithms to assess the complex cellular heterogeneity within the failing myocardium. Despite variations in the underlying logic and weights of the different immune infiltration analysis algorithms, all five methods consistently indicated an upregulated expression of T cells or their subgroups in the failing myocardium as compared to normal myocardium. Additionally, at least two of these algorithms support the presence of differences in Mast cells, B cells, NK cells, and macrophages within the failing myocardium. Hence, the results of our immune infiltration analysis suggest alterations in certain immune cell levels in the failing myocardium when contrasted with normal myocardium.

In order to investigate whether the altered levels of immune cells are causally associated with the onset and progression of HF, we systematically provided genetic support for the causal relationship between 731 immune cell phenotypes across seven groups of immune cells and HF through bidirectional MR analysis. We observed a causal relationship between 31 immune cell phenotypes in B cells, cDC, TBNK, Treg, Myeloid cells and Maturation stages of T cells with HF in forward MR. In contrast, in reverse MR, there was no evidence to support a causal relationship between HF and 731 immune cell phenotypes. There was no bidirectional causality between HF and 731 immune cell phenotypes. As the results of the reverse MR analysis do not support a causal relationship between HF and immune cells, this suggests that the changes in immune cell levels observed in the failing myocardium are not caused by heart failure itself. Although the reasons for these alterations in immune cell expression are not yet clear, they may have an impact on the progression of heart failure.

The results of the immune infiltration analysis indicate an increased expression of T cells and their subgroups in the failing myocardium. However, the MR analysis reveals that the impact of different immune phenotypes within T cell subgroups, such as CD4 or CD8, on heart failure is complex, which may account for the discouraging outcomes of immune modulatory therapies. Alba Vilella-Figuerola et al. demonstrated that activated CD45+ T cells were positively correlated with the severity of congestive HF, which was consistent with our MR analysis of CD45 on lymphocyte positively correlated with HF^34^. Elisa Martini et al. focused their research on CD45+ cells and found that activated immune cells were strongly associated with HF^35^. Yun-Long Zhang et al. concluded that targeting CD11b was associated with cardiac remodeling and may benefit HF^36^. Animal studies have demonstrated that the systemic activation of induced regulatory T cells can be employed in anti-remodeling therapy for the repair of ischemic tissues^37^. The expression of CD69 on regulatory T cells can also protect against immune injury post-myocardial infarction, thereby preventing heart failure^38^. Furthermore, research has indicated that cardiac fibroblasts expressing xenogeneic antigens can be effectively targeted and eliminated through the adoptive transfer of antigen-specific CD8+ T cells, which may alleviate myocardial fibrosis^39^. Therefore, immunomodulatory therapies that intervene with T cells are promising, but it is important to consider the regulation of specific subgroups^40^.

MR results identified 16 immune cell phenotypes in B cells that were significantly associated with HF. B cells are a key part of the organism’s immune system, participating in the immune response through the production of antibodies, as well as in regulating the activities of other immune cells^41^. B cells regulate macrophage phenotype in the myocardium and play an important role in maintaining myocardial homeostasis^42^. Several studies have shown that B cells can influence cardiac remodeling and inflammatory responses after myocardial infarction, thereby mediating HF^43,44^. In addition, antibodies may also mediate myocardial injury through complement-associated mechanisms and may interact with cardiomyocytes or other immune cells to affect myocardial function^45–47^. Andrea M. Cordero-Reyes et al. concluded that pathways of B cell activation were intimately involved in the progression of HF^48^.

MR results identified 5 immune cell phenotypes in TBNK cells that were significantly associated with HF. Existing studies suggest that natural killer (NK) cells contribute to the mitigation of cardiac inflammation and fibrosis by inhibiting the infiltration of eosinophils, thereby exerting an anti-heart failure effect^49^. Additionally, NK cells may offer protective benefits in the aftermath of a heart attack by boosting the expression of cardioprotective cytokines, including IL-10, which aids in the remodeling and preservation of left ventricular function^50^. In a contradiction, findings derived from the xCell algorithm indicate a reduction in NK cell expression within failing myocardial tissue, which is at odds with the results yielded by the ssGSEA and QuanTIseq algorithms. This is further complicated by a clinical study involving 82 patients with end-stage heart failure, which revealed that the levels of circulating NK cells in patients were lower compared to healthy individuals^51^.

Our study also identified certain immune cell phenotypes that exhibit causal relationships with HF, yet to date, there have been no clinical studies or reports on their association with HF. The role of these immune cell phenotypes in the pathogenesis of heart failure holds promise and warrants further investigation. Additionally, our bidirectional Mendelian randomization does not support a causal relationship between HF and 731 immune cell phenotypes. This suggests that certain immune cell phenotypes that could potentially ameliorate or accelerate the course of heart failure are not influenced by the progression of the condition.

In this study, we initially confirmed the levels of immune cells in failing myocardium using five distinct immune infiltration analysis algorithms. Subsequently, a two-sample bidirectional MR analysis was performed, which allowed for large sample sizes and high statistical efficiency. The conclusions of the MR analysis in this research are based on exploring the causal relationship between the two variables with genetic instrumental variables, and causal inference and result verification were carried out using various MR analysis methods. Therefore, the results are robust and not affected or confounded by pleiotropy at horizontal levels.

However, our study should consider some limitations. Firstly, due to the differing logic and weights of the immune infiltration analysis algorithms used, there are inconsistencies in the computed results, which require further experimental support. Secondly, we selected IVs with a P-value threshold of P < 1E-5, therefore, the strength of the associations of these IVs may not be robust, despite allowing for a more comprehensive assessment of the association between immune cell phenotypes and heart failure. Thirdly, as this study is based on a European population, the applicability of the results to other ethnic groups remains debatable, limiting the breadth of our results. Additionally, despite multiple sensitivity analyses, the assessment of horizontal pleiotropy remains limited. While statistically it is considered satisfactory to eliminate heterogeneity and horizontal pleiotropy, it cannot be fully guaranteed that such factors are absent in the clinical environment. Finally, comprehensive clinical trials are needed to draw clinical conclusions. Therefore, to elucidate the association and mechanisms between individual immune cell phenotypes and heart failure, we need more extensive GEO and GWAS databases, as well as further analytical methods or experimental validations.

## CONCLUSION

We determined the levels of immune cells in failing myocardium using five immune infiltration analysis algorithms and demonstrated the causal relationships between various immune cell phenotypes and heart failure through comprehensive bidirectional two-sample MR analysis. This opens new avenues for researchers to explore the biological mechanisms of heart failure. More importantly, it offers innovative perspectives for investigating immunocyte therapies for heart failure, which may aid in the prevention and treatment of heart failure.

## Data Availability

Publicly available datasets were analyzed in this study. This data can be found here: GSE57338 and GWAS Catalog (https://www.ebi.ac.uk/gwas/downloads/summary-statistics)

## Acknowledgements

Not applicable.

## CRediT authorship contribution statement

Yiding Yu: Conceptualization, Methodology, Validation, Formal analysis, Investigation, Resources, Visualization, Writing – original draft, Writing – review & editing. Huajing Yuan: Methodology, Validation, Formal analysis, Software, Data curation, Writing – original draft. Wenwen Liu: Software, Data curation, Writing – review & editing. Yitao Xue: Supervision, Funding acquisition, Writing – review & editing. Xiujuan Liu: Project administration, Supervision, Writing – review & editing. Wangjun Hou: Project administration, Writing – review & editing, Funding acquisition. Yan Li: Project administration, Writing – review & editing, Funding acquisition.

## Consent for publication

Not applicable.

## Ethics approval and consent to participate

Not applicable.

## REFERENCES

1. S.A. Hunt, W.T. Abraham, M.H. Chin, A.M. Feldman, G.S. Francis, T.G. Ganiats, M. Jessup, M.A. Konstam, D.M. Mancini, K. Michl, J.A. Oates, P.S. Rahko,M.A. Silver, L.W. Stevenson, C.W. Yancy, E.M. Antman, S.C. Smith Jr., C.D. Adams,J.L. Anderson, D.P. Faxon, V. Fuster, J.L. Halperin, L.F. Hiratzka, S.A. Hunt,A.K. Jacobs, R. Nishimura, J.P. Ornato, R.L. Page, B. Riegel, Acc/aha 2005 guideline update for the diagnosis and management of chronic heart failure in the adult summary article, J. Am. Coll. Cardiol. 46 (2005) 1116–1143, 10.1016/j.jacc2005.08023.

2. Savarese, G., Becher, P. M., Lund, L. H., Seferovic, P., Rosano, G. M. C., & Coats, A. J. S. (2023). Global burden of heart failure: a comprehensive and updated review of epidemiology. Cardiovascular research, 118(17), 3272–3287. 10.1093/cvr/cvac013

3. Roger V. L. (2021). Epidemiology of Heart Failure: A Contemporary Perspective. Circulation research, 128(10), 1421–1434. 10.1161/CIRCRESAHA.121.318172

4. Towbin, J., & Bowles, N. (2002). The failing heart. Nature, 415, 227–233. 10.1038/415227a.

5. Lymperopoulos, A., Rengo, G., & Koch, W. (2013). Adrenergic Nervous System in Heart Failure: Pathophysiology and Therapy. Circulation Research, 113, 739–753. 10.1161/CIRCRESAHA.113.300308.

6. Ventura-clapier, R., Garnier, A., & Veksler, V. (2004). Energy metabolism in heart failure. The Journal of Physiology, 555. 10.1113/jphysiol.2003.055095.

7. Kemp, C., & Conte, J. (2012). The pathophysiology of heart failure.. Cardiovascular pathology : the official journal of the Society for Cardiovascular Pathology, 21 5, 365–71 . 10.1016/j.carpath.2011.11.007.

8. Slørdal, L., & Spigset, O. (2006). Heart Failure Induced by Non-Cardiac Drugs. Drug Safety, 29, 567–586. 10.2165/00002018-200629070-00003.

9. Oka, T., & Komuro, I. (2008). Molecular mechanisms underlying the transition of cardiac hypertrophy to heart failure.. Circulation journal : official journal of the Japanese Circulation Society, 72 Suppl A, A13-6 . 10.1253/CIRCJ.CJ-08-0481.

10. Papait, R., Greco, C., Kunderfranco, P., Latronico, M., & Condorelli, G. (2013). Epigenetics: a new mechanism of regulation of heart failure?. Basic Research in Cardiology, 108. 10.1007/s00395-013-0361-1.

11. Swirski, F., & Nahrendorf, M. (2018). Cardioimmunology: the immune system in cardiac homeostasis and disease. Nature Reviews Immunology, 18, 733–744. 10.1038/s41577-018-0065-8.

12. Peet, C., Ivetic, A., Bromage, D. I., & Shah, A. M. (2020). Cardiac monocytes and macrophages after myocardial infarction. Cardiovascular research, 116(6), 1101–1112. 10.1093/cvr/cvz336

13. Santos-Zas, I., Lemarié, J., Zlatanova, I., Cachanado, M., Seghezzi, J. C., Benamer, H., Goube, P., Vandestienne, M., Cohen, R., Ezzo, M., Duval, V., Zhang, Y., Su, J. B., Bizé, A., Sambin, L., Bonnin, P., Branchereau, M., Heymes, C., Tanchot, C., Vilar, J., … Ait-Oufella, H. (2021). Cytotoxic CD8+ T cells promote granzyme B-dependent adverse post-ischemic cardiac remodeling. Nature communications, 12(1), 1483. 10.1038/s41467-021-21737-9

14. Vdovenko, D., & Eriksson, U. (2018). Regulatory Role of CD4+ T Cells in Myocarditis. Journal of immunology research, 2018, 4396351. 10.1155/2018/4396351

15. Bermea, K., Bhalodia, A., Huff, A., Rousseau, S., & Adamo, L. (2022). The Role of B Cells in Cardiomyopathy and Heart Failure. Current cardiology reports, 24(8), 935–946. 10.1007/s11886-022-01722-4

16. Lu, Y., Xia, N., & Cheng, X. (2021). Regulatory T Cells in Chronic Heart Failure. Frontiers in immunology, 12, 732794. 10.3389/fimmu.2021.732794

17. Tang, X., Wang, P., Zhang, R., Watanabe, I., Chang, E., Vinayachandran, V., Nayak, L., Lapping, S., Liao, S., Madera, A., Sweet, D. R., Luo, J., Fei, J., Jeong, H. W., Adams, R. H., Zhang, T., Liao, X., & Jain, M. K. (2022). KLF2 regulates neutrophil activation and thrombosis in cardiac hypertrophy and heart failure progression. The Journal of clinical investigation, 132(3), e147191. 10.1172/JCI147191

18. Saleh, D., Jones, R. T. L., Schroth, S. L., Thorp, E. B., & Feinstein, M. J. (2023). Emerging Roles for Dendritic Cells in Heart Failure. Biomolecules, 13(10), 1535. 10.3390/biom13101535

19. Ong, S., Rose, N., & Čiháková, D. (2017). Natural killer cells in inflammatory heart disease.. Clinical immunology, 175, 26–33 . 10.1016/j.clim.2016.11.010.

20. Fildes, J. E., Shaw, S. M., Yonan, N., & Williams, S. G. (2009). The immune system and chronic heart failure: is the heart in control?. Journal of the American College of Cardiology, 53(12), 1013–1020. 10.1016/j.jacc.2008.11.046

21. Liu, Y., Morley, M., Brandimarto, J., Hannenhalli, S., Hu, Y., Ashley, E. A., Tang, W. H., Moravec, C. S., Margulies, K. B., Cappola, T. P., Li, M., & MAGNet consortium (2015). RNA-Seq identifies novel myocardial gene expression signatures of heart failure. Genomics, 105(2), 83–89. 10.1016/j.ygeno.2014.12.002

22. Newman, A. M., Liu, C. L., Green, M. R., Gentles, A. J., Feng, W., Xu, Y., Hoang, C. D., Diehn, M., & Alizadeh, A. A. (2015). Robust enumeration of cell subsets from tissue expression profiles. Nature methods, 12(5), 453–457. 10.1038/nmeth.3337

23. Aran, D., Hu, Z., & Butte, A. J. (2017). xCell: digitally portraying the tissue cellular heterogeneity landscape. Genome biology, 18(1), 220. 10.1186/s13059-017-1349-1

24. Bindea, G., Mlecnik, B., Tosolini, M., Kirilovsky, A., Waldner, M., Obenauf, A. C., Angell, H., Fredriksen, T., Lafontaine, L., Berger, A., Bruneval, P., Fridman, W. H., Becker, C., Pagès, F., Speicher, M. R., Trajanoski, Z., & Galon, J. (2013). Spatiotemporal dynamics of intratumoral immune cells reveal the immune landscape in human cancer. Immunity, 39(4), 782–795. 10.1016/j.immuni.2013.10.003

25. 25. Becht, E., Giraldo, N. A., Lacroix, L., Buttard, B., Elarouci, N., Petitprez, F., Selves, J., Laurent-Puig, P., Sautès-Fridman, C., Fridman, W. H., & de Reyniès, A. (2016). Estimating the population abundance of tissue-infiltrating immune and stromal cell populations using gene expression. Genome biology, 17(1), 218. 10.1186/s13059-016-1070-5

26. Finotello, F., Mayer, C., Plattner, C., Laschober, G., Rieder, D., Hackl, H., Krogsdam, A., Loncova, Z., Posch, W., Wilflingseder, D., Sopper, S., Ijsselsteijn, M., Brouwer, T. P., Johnson, D., Xu, Y., Wang, Y., Sanders, M. E., Estrada, M. V., Ericsson- Gonzalez, P., Charoentong, P., … Trajanoski, Z. (2019). Molecular and pharmacological modulators of the tumor immune contexture revealed by deconvolution of RNA-seq data. Genome medicine, 11(1), 34. 10.1186/s13073-019-0638-6

27. Bowden, J., Davey Smith, G., Haycock, P. C., & Burgess, S. (2016). Consistent Estimation in Mendelian Randomization with Some Invalid Instruments Using a Weighted Median Estimator. Genetic epidemiology, 40(4), 304–314. 10.1002/gepi.21965

28. Orrù, V., Steri, M., Sidore, C., Marongiu, M., Serra, V., Olla, S., Sole, G., Lai, S., Dei, M., Mulas, A., Virdis, F., Piras, M. G., Lobina, M., Marongiu, M., Pitzalis, M., Deidda, F., Loizedda, A., Onano, S., Zoledziewska, M., Sawcer, S., … Cucca, F. (2020). Complex genetic signatures in immune cells underlie autoimmunity and inform therapy. Nature genetics, 52(10), 1036–1045. 10.1038/s41588-020-0684-4

29. Shah, S., Henry, A., Roselli, C., Lin, H., Sveinbjörnsson, G., Fatemifar, G., Hedman, Å. K., Wilk, J. B., Morley, M. P., Chaffin, M. D., Helgadottir, A., Verweij, N., Dehghan, A., Almgren, P., Andersson, C., Aragam, K. G., Ärnlöv, J., Backman, J. D., Biggs, M. L., Bloom, H. L., … Lumbers, R. T. (2020). Genome-wide association and Mendelian randomisation analysis provide insights into the pathogenesis of heart failure. Nature communications, 11(1), 163. 10.1038/s41467-019-13690-5

30. Li, P., Wang, H., Guo, L., Gou, X., Chen, G., Lin, D., Fan, D., Guo, X., & Liu, Z. (2022). Association between gut microbiota and preeclampsia-eclampsia: a two-sample Mendelian randomization study. BMC medicine, 20(1), 443. 10.1186/s12916-022-02657-x

31. Hemani, G., Zheng, J., Elsworth, B., Wade, K. H., Haberland, V., Baird, D., Laurin, C., Burgess, S., Bowden, J., Langdon, R., Tan, V. Y., Yarmolinsky, J., Shihab, H. A., Timpson, N. J., Evans, D. M., Relton, C., Martin, R. M., Davey Smith, G., Gaunt, T. R., & Haycock, P. C. (2018). The MR-Base platform supports systematic causal inference across the human phenome. eLife, 7, e34408. 10.7554/eLife.34408

32. Bowden, J., Davey Smith, G., & Burgess, S. (2015). Mendelian randomization with invalid instruments: effect estimation and bias detection through Egger regression. International journal of epidemiology, 44(2), 512–525. 10.1093/ije/dyv080

33. Verbanck, M., Chen, C. Y., Neale, B., & Do, R. (2018). Detection of widespread horizontal pleiotropy in causal relationships inferred from Mendelian randomization between complex traits and diseases. Nature genetics, 50(5), 693–698. 10.1038/s41588-018-0099-7

34. Vilella-Figuerola A, Padro T, Roig E, Mirabet S, Badimon L. New factors in heart failure pathophysiology: Immunity cells release of extracellular vesicles. Front Cardiovasc Med (2022) 9: 939625. doi:10.3389/fcvm.2022.939625

35. Martini E, Kunderfranco P, Peano C, Carullo P, Cremonesi M, Schorn T, et al. Single-Cell Sequencing of Mouse Heart Immune Infiltrate in Pressure Overload-Driven Heart Failure Reveals Extent of Immune Activation. Circulation (2019) 140: 2089–107. doi:10.1161/CIRCULATIONAHA.119.041694

36. Zhang YL, Bai J, Yu WJ, Lin QY, Li HH. CD11b mediates hypertensive cardiac remodeling by regulating macrophage infiltration and polarization. J Adv Res (2024) 55: 17–31. doi:10.1016/j.jare.2023.02.010

37. Choo EH, Lee JH, Park EH, Park HE, Jung NC, Kim TH, Koh YS, Kim E, Seung KB, Park C, Hong KS, Kang K, Song JY, Seo HG, Lim DS, Chang K. Infarcted Myocardium-Primed Dendritic Cells Improve Remodeling and Cardiac Function After Myocardial Infarction by Modulating the Regulatory T Cell and Macrophage Polarization. Circulation. 2017 Apr 11;135(15):1444–1457. doi: 10.1161/CIRCULATIONAHA.116.023106.

38. Blanco-Domínguez R, de la Fuente H, Rodríguez C, Martín-Aguado L, Sánchez-Díaz R, Jiménez-Alejandre R, Rodríguez-Arabaolaza I, Curtabbi A, García-Guimaraes MM, Vera A, Rivero F, Cuesta J, Jiménez-Borreguero LJ, Cecconi A, Duran-Cambra A, Taurón M, Alonso J, Bueno H, Villalba-Orero M, Enríquez JA, Robson SC, Alfonso F, Sánchez-Madrid F, Martínez-González J, Martín P. CD69 expression on regulatory T cells protects from immune damage after myocardial infarction. J Clin Invest. 2022 Nov 1;132(21):e152418. doi: 10.1172/JCI152418.

39. Aghajanian H, Kimura T, Rurik JG, Hancock AS, Leibowitz MS, Li L, Scholler J, Monslow J, Lo A, Han W, Wang T, Bedi K, Morley MP, Linares Saldana RA, Bolar NA, McDaid K, Assenmacher CA, Smith CL, Wirth D, June CH, Margulies KB, Jain R, Puré E, Albelda SM, Epstein JA. Targeting cardiac fibrosis with engineered T cells. Nature. 2019 Sep;573(7774):430-433. doi: 10.1038/s41586-019-1546-z.

40. Lu Y, Xia N, Cheng X. Regulatory T Cells in Chronic Heart Failure. Front Immunol. 2021 Sep 22;12:732794. doi: 10.3389/fimmu.2021.732794.

41. Bermea K, Bhalodia A, Huff A, Rousseau S, Adamo L. The Role of B Cells in Cardiomyopathy and Heart Failure. Curr Cardiol Rep (2022) 24: 935–46. doi:10.1007/s11886-022-01722-4

42. Rocha-Resende C, Pani F, Adamo L. B cells modulate the expression of MHC-II on cardiac CCR2(-) macrophages. J Mol Cell Cardiol (2021) 157: 98–103. doi:10.1016/j.yjmcc.2021.05.003

43. Heinrichs M, Ashour D, Siegel J, Buchner L, Wedekind G, Heinze KG, et al. The healing myocardium mobilizes a distinct B-cell subset through a CXCL13-CXCR5- dependent mechanism. Cardiovasc Res (2021) 117: 2664–76. doi:10.1093/cvr/cvab181

44. Yan X, Anzai A, Katsumata Y, Matsuhashi T, Ito K, Endo J, et al. Temporal dynamics of cardiac immune cell accumulation following acute myocardial infarction. J Mol Cell Cardiol (2013) 62: 24–35. doi:10.1016/j.yjmcc.2013.04.023

45. Zhang M, Michael LH, Grosjean SA, Kelly RA, Carroll MC, Entman ML. The role of natural IgM in myocardial ischemia-reperfusion injury. J Mol Cell Cardiol (2006) 41: 62–67. doi:10.1016/j.yjmcc.2006.02.006

46. Ludwig RJ, Vanhoorelbeke K, Leypoldt F, Kaya Z, Bieber K, McLachlan SM, et al. Mechanisms of Autoantibody-Induced Pathology. Front Immunol (2017) 8: 603. doi:10.3389/fimmu.2017.00603

47. Haudek SB, Trial J, Xia Y, Gupta D, Pilling D, Entman ML. Fc receptor engagement mediates differentiation of cardiac fibroblast precursor cells. Proc Natl Acad Sci U S A (2008) 105: 10179–84. doi:10.1073/pnas.0804910105

48. Cordero-Reyes AM, Youker KA, Torre-Amione G. The role of B-cells in heart failure. Methodist Debakey Cardiovasc J (2013) 9: 15–19. doi:10.14797/mdcj-9-1-15

49. Ong S, Ligons DL, Barin JG, Wu L, Talor MV, Diny N, Fontes JA, Gebremariam E, Kass DA, Rose NR, Čiháková D. Natural killer cells limit cardiac inflammation and fibrosis by halting eosinophil infiltration. Am J Pathol. 2015 Mar;185(3):847–61. doi: 10.1016/j.ajpath.2014.11.023.

50. Sobirin MA, Kinugawa S, Takahashi M, Fukushima A, Homma T, Ono T, Hirabayashi K, Suga T, Azalia P, Takada S, Taniguchi M, Nakayama T, Ishimori N, Iwabuchi K, Tsutsui H. Activation of natural killer T cells ameliorates postinfarct cardiac remodeling and failure in mice. Circ Res. 2012 Sep 28;111(8):1037–47. doi: 10.1161/CIRCRESAHA.112.270132.

51. Vredevoe DL, Widawski M, Fonarow GC, Hamilton M, Martínez-Maza O, Gage JR. Interleukin-6 (IL-6) expression and natural killer (NK) cell dysfunction and anergy in heart failure. Am J Cardiol. 2004 Apr 15;93(8):1007–11. doi: 10.1016/j.amjcard.2003.12.054.

